# “Till How Long Will We Struggle?”: A qualitative exploration of the lingering mental toll of COVID-19 on young people in Sudan and Malawi

**DOI:** 10.1101/2025.09.14.25335714

**Authors:** Shahd Mekki, Sandra Jumbe

**Affiliations:** Wolfson Institute of Population Health, Queen Mary University of London, London, United Kingdom; Department of Research and Corporate Governance, Millennium University, Blantyre, Malawi

## Abstract

During the COVID-19 pandemic, various aspects of life were **s**ignificantly disrupted due to ill health, economic instability, and reduced social interactions precipitated by pandemic related restrictions. These disruptions exacerbated stress among young populations across the globe. This qualitative study explores how the pandemic affected the mental health of young Africans aged 18 to 24 years residing in Malawi and Sudan.

We conducted six focus group discussions (FGDs), with 25 participants from Malawi and Sudan. Participants were recruited through a poster posted on Facebook and X (previously Twitter). These FGDs were recorded, transcribed verbatim, and analysed using thematic analysis.

Three key themes emerged from the analysis: (1) main challenges facing youth, including economic hardship, social pressures, and limited educational and employment opportunities; (2) stigma, lack of awareness, and limited availability of trained professionals as barriers to accessing mental health services; and (3) recommendations for improving mental health service delivery. Across these themes, participants voiced a need for community outreach, integrating mental health into primary care services, and providing culturally sensitive support tailored to the needs of young people.

This study highlights key mental health challenges faced by young people in low-resource settings during the COVID-19 pandemic, emphasising the need for tailored policies to support youth mental health during and after pandemics. Results underscore the need for community-based approaches to enhance mental health literacy and develop youth-focused mental health services in Malawi and Sudan. This research also provides valuable insights into potential preventive measures and interventions needed to mitigate these effects.

## Introduction

The global impact of the coronavirus outbreak in late 2019 fundamentally reshaped lives, economies, and healthcare systems worldwide. COVID-19 swiftly escalated into a global health crisis, prompting widespread concern for health and stringent quarantine measures. In January 2020, the World Health Organization (WHO) declared COVID-19 a global public health emergency, underscoring its severity and the urgent need for coordinated global responses [1].

Originating as severe acute respiratory syndrome coronavirus (SARS-CoV-2) viral pneumonia, COVID-19 rapidly evolved into a global pandemic as it spread efficiently. This unprecedented catastrophe not only increased mortality rates but also exacerbated mental health challenges worldwide [2]. Historical infectious diseases like H1N1 and SARS serve as previous examples of threats to global mental well-being, often resulting in conditions like post-traumatic stress disorder (PTSD) [3].

Mental health is pivotal to how individuals navigate life’s challenges, regulate emotions, and foster healthy relationships. The COVID-19 pandemic exacerbated mental health disparities, particularly among young people aged 15-24 years old, with studies revealing alarmingly high rates of anxiety and depression globally [4, 5]. Factors such as social distancing measures, school closures, and economic instability further compounded the mental health struggles of youth. The intersection of these stressors created complex and multifaceted negative impacts on mental health that are difficult to quantify.

Navigating the mental health landscape during the COVID-19 pandemic was challenging. While some advocated for lockdowns as essential tools in combating the virus, others raised concerns about their unintended consequences, including worsened mental health outcomes [6]. Additionally, issues of equity and social justice in mental health provision came to the forefront, highlighting existing disparities in access to care and resources [7]. The pandemic laid bare and exacerbated pre-existing global socioeconomic inequalities that intersect with healthcare, disproportionately affecting vulnerable populations such as women, ethnic minorities, and underserved communities in low– and middle-income countries (LMICs) [8]. In regions like Africa, the pandemic intensified pre-existing challenges like economic disparities, limited healthcare access, and governmental fragility [9]. Despite the extensive discourse on global mental health, limited research infrastructure and resources in LMICs hinder efforts to understand the prevalence and consequences of mental health issues, highlighting gaps in mental health literacy and access to support services [10]. This gap in knowledge is particularly concerning given the unique sociocultural contexts of these regions, which directly impacts management of mental health issues.

There is also a significant research gap regarding the mental health experiences of young people in LMICs, especially among African populations. The scarcity of research on the mental health experiences of young people in Africa poses significant challenges to developing effective, context-specific interventions. In sub-Saharan Africa, studies reveal high prevalence rates of mental health issues among adolescents—27% for depression, 30% for anxiety disorders, and 41% for emotional and behavioural problems—yet mental health services remain severely under-resourced, with only 13 out of 48 countries on the continent having standalone adolescent mental health policies and treatment gaps of up to 90% [11, 12].

This research aimed to bridge the gap in understanding the effects of the COVID-19 pandemic on the mental health of young people in Malawi and Sudan, two countries with distinct sociocultural contexts. It formed part of a larger research project titled the *Implementation of a Mental Health Literacy e-Curriculum in Malawi Universities* (MHLeC) study—a 4-year programme designed to increase mental health literacy among African populations through educational settings [17]. We started the project with community engagement work, engaging with groups of young people from these two countries as key stakeholders to understand how COVID-19 had impacted them and their peers’ mental health accounting for contextual variation [18]. Community engagement (CE) is an important part of health research practice particularly in low-income settings or with disadvantaged groups [19, 20]. We prioritised partnership through CE with people with lived experience throughout the research process to help us identify problems raised by those directly affected (our stakeholders), and learn from them regarding ideas on potential solutions, ensuring that our research was relevant [20]. We report findings from focus groups with these youth groups in Malawi conducted as part of CE. Using focus group discussions, we explored how young people navigated the psychological challenges posed by the pandemic and identified both structural and personal factors influencing their mental well-being. We specifically sought to explore young people’s lives changed, the challenges they faced, and adaptations they made to the pandemic phenomenon with a focal lens on mental health. Our goal was to use in-depth qualitative analysis to elucidate underlying dynamics and understanding of the mental health challenges faced by the population to inform identification of culturally relevant mental health support strategies.

## Materials and Methods

### Study Design

We used an exploratory qualitative approach to investigate the mental health impact of the COVID-19 pandemic on young people from Malawi and Sudan. FGDs were selected as the primary data collection method due to their effectiveness in generating rich, nuanced data through social interaction and collective sense-making among participants. As Acocella noted, FGDs offer distinct advantages over other methods due to their innovative, cost-effective, and easily organised nature [13].

Participants in focus groups often engage in mature interactions, facilitating the collection of relevant information within a limited timeframe whilst swiftly unveiling collective opinions.

### Participant Recruitment and Sampling

25 participants aged between 18 and 24 years who self-identify as Malawian or Sudanese were purposively recruited to capture diverse perspectives and lived experiences of how the COVID-19 pandemic affected the mental health of young people in their respective communities.

As this study was conducted during the COVID-19 pandemic, face to face research activities had been halted. Recruitment was mainly done through social media platforms—Facebook and X (formerly Twitter)—using digital posters distributed through university societies, youth-led organisations, and community networks in the two countries (Figure 1). Purposeful sampling was also used to facilitate recruitment – specifically, individuals who expressed an interest to take part in the study after seeing our posters on social media were asked to promote the study among their existing peer networks.

**Figure 1:**
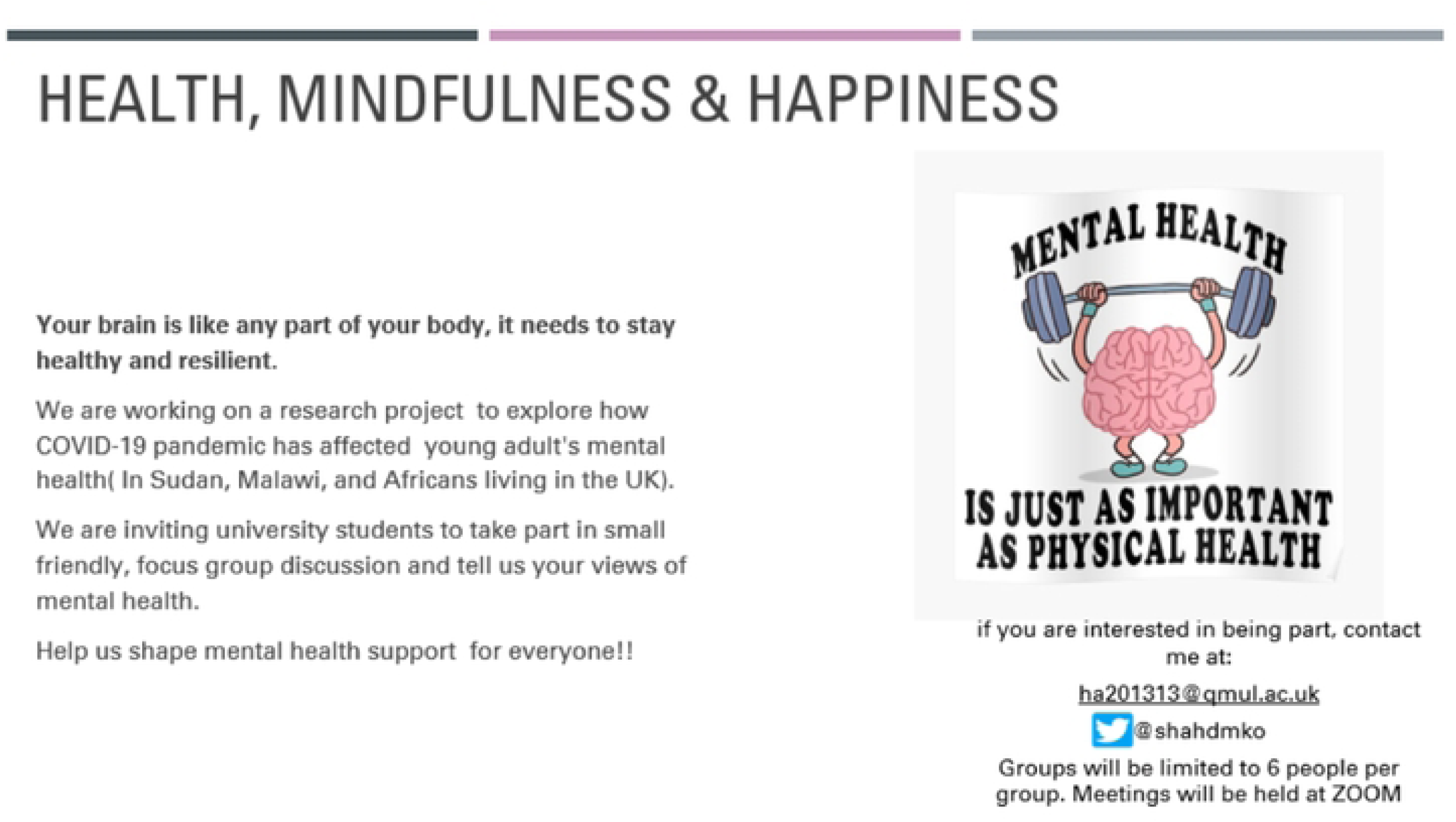
Study poster.

Participants received a study information sheet and were given the opportunity to ask questions before their scheduled focus groups.

### Data Collection

Study recruitment began 8^th^ March 2021 when the study poster was posted on social media. FGDs were conducted virtually using Zoom between March and 6^th^ May 2021, each lasting approximately two hours. Before each session, participants provided verbal informed consent, which was recorded. Demographic data, including age, gender, and nationality, were collected to help contextualise findings. Each session began with a short introductory icebreaker to foster rapport and create a comfortable, respectful environment for discussion.

To reduce language-related barriers, participants were given the option to speak in English and/or their native language (Arabic or Chichewa). This flexibility ensured that participants could express themselves fully and authentically. Arabic was translated by the lead researcher (Shahd Mekki), and Chichewa was translated by Dr Sandra Jumbe (research supervisor) both fluent in these respective languages and English.

FGDs were led by both authors, who took a semi-structured approach facilitated by a topic guide across all FGDs to ensure consistency in questions posed (Figure 2).

**Figure 2:**
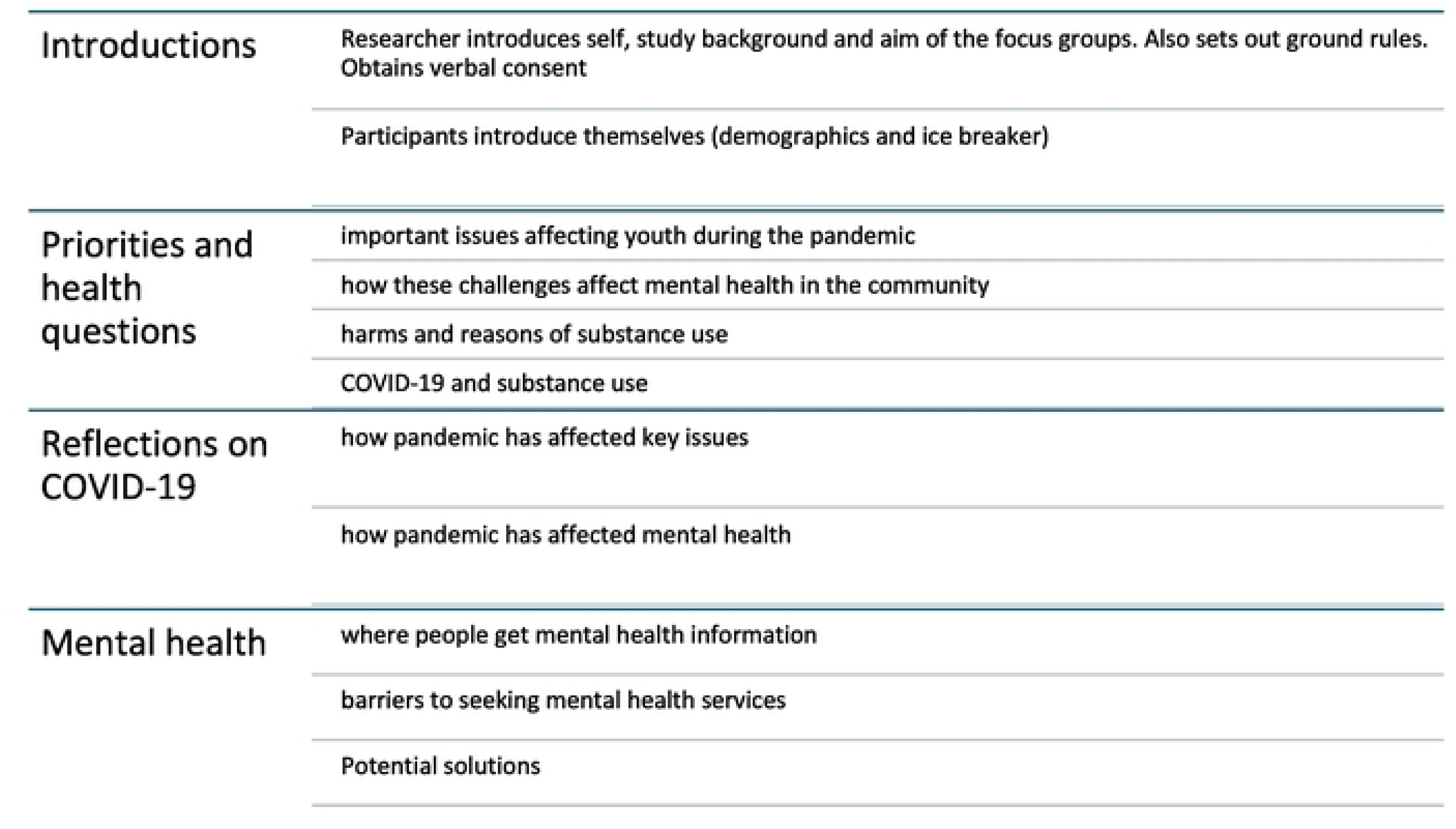
Summarised topic guide for community youth stakeholders’ focus groups.

### Data Management and Analysis

Audio recordings of the FGDs were securely stored and transcribed verbatim by the lead researcher. Transcripts were anonymised, checked for accuracy, and reviewed by the research supervisor to ensure data integrity. The transcripts were imported into qualitative analysis software, NVivo.

Thematic analysis was employed to explore patterns and insights within the data [16]. This process involved multiple steps: (1) familiarisation with the data through repeated readings, (2) initial line-by-line inductive coding, (3) grouping similar codes into broader categories, (4) identification and refinement of key themes and subthemes, and (5) naming and defining each theme. Themes were derived from the data itself, rather than predefined categories, reflecting participants’ own words and lived experiences.

To support the credibility and trustworthiness of the findings, peer debriefing and intercoder discussion were employed during analysis. Discrepancies in interpretation were resolved through dialogue and consensus between the two researchers. Representative quotes were selected to illustrate each theme and ensure that participants’ voices were authentically represented.

### Reflexivity

Throughout the research process, the authors engaged in continuous reflexivity, acknowledging their own positionalities as researchers of African origin with personal and professional interest in young people’s mental health. Field notes and memos were maintained to document observations, reactions, and potential biases. These were revisited during analysis to ensure that findings were grounded in participants’ perspectives rather than researchers’ assumptions.

### Ethical Considerations

Ethical approval was obtained from of the Queen Mary Ethics of Research Committee on 14th December 2020, following the review of the study protocol and associated materials (Reference: QMERC2424a), before commencing the study. All participants received verbal and written information about the study prior to giving their informed consent. They were assured of confidentiality and the voluntary nature of their involvement. Identifiable data were anonymised, and data storage complied with institutional data protection policies.

## Results

### Participants’ characteristics

Initially, thirty individuals were recruited to the study. However, due to technical difficulties such as poor internet connectivity and limited access to electricity, five were unable to join their assigned focus groups and subsequently chose not to continue their participation in the research. Demographic details of the participants are presented in Table 1. There was a similar number of participants between Malawi and Sudan. The sample was predominantly female (n=23).

**Table 1.** – Characteristics of Participants.

|  |  |  |
| --- | --- | --- |
| **Demographics (n = 25)** |  |  |
| **Age range (years)** |  | 18-24 years |
| **Gender** |  |  |
| Females | 23 |  |
| Males |  | 2 |
| **Country of origin** |  |  |
| Sudan |  | 12 |
| Malawi | 13 |  |

### Overview of themes

Following thematic analysis of focus group discussions (FGDs) on the impact of the COVID-19 pandemic on the mental health of young people in Malawi and Sudan, three central themes were identified: (1) Main challenges affecting young people’s mental health, (2) Barriers to accessing mental health services—primarily related to social and financial obstacles, and (3) Ways to and access mental health support. Participants consistently reported a decline in their mental wellbeing, citing increased anxiety, depression, and stress, which were intensified by the uncertainty and isolation resulting from quarantine measures and social distancing during the pandemic. They also highlighted that the disruption of daily routines and limited access to support networks further compounded their mental health struggles.

The overarching theme among participants as they recounted their lived experiences during the COVID-19 pandemic was that of a never-ending uncertainty which brought on persisting mental health struggle. Participants highlighted additional stress from academic or career-related disturbances, leading to feelings of despondency and powerlessness. Substance abuse was noted as a significant issue, with young people turning to alcohol and drugs to cope with the stress and anxiety induced by the pandemic. This created a vicious cycle of addiction and further deteriorated mental health.

Economic instability, including job losses and business closures, was a prominent concern, particularly for fresh graduates and early-career individuals in both Malawi and Sudan. Social distancing, online learning, and lockdowns exacerbated loneliness and social isolation, with participants reporting increased mood disorder symptoms due to separation from friends, family, and community. The lack of social engagement and support was particularly evident.

Stigma was a critical obstacle to seeking mental health services across both country settings, where negative cultural biases and attitudes towards mental illness were prevalent. This hindered participants from talking about their mental health struggles and seeking support. Participants from both countries also faced challenges to access mental health professionals and resources.

Figure 3 represents these key themes and associated subthemes. Illustrative quotes are provided throughout the results section, with quotes ending in *[M-FG]* from Malawian participants and *[S-FG]* from Sudanese participants.

**Figure 3.**
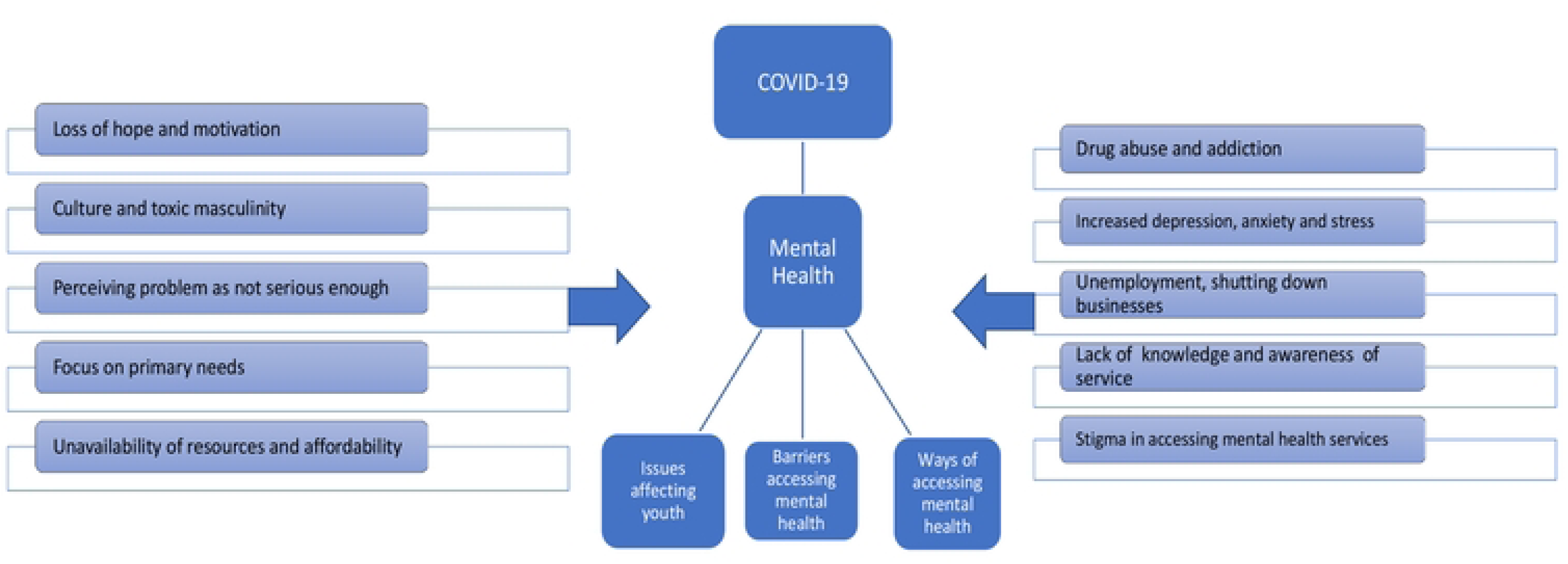
– thematic map.

### Theme 1: Main challenges affecting young people’s mental health

Table 2 provides a summary of key topics raised along with their frequency within and across FGDs.as participants talked about important issues that are affecting young people in their respective countries.

**Table 2.** – Topics linked to ‘Main challenges Affecting Youth mental health’.

|  | **<u>Sudan (3)</u>** | **<u>Malawi (3)</u>** | **<u>Total FGD (6)</u>** |
| --- | --- | --- | --- |
| <u>Substance abuse and addiction</u> | <u>3</u> | <u>3</u> | <u>6</u> |
| <u>Unemployment and businesses shutting down</u> | <u>2</u> | <u>3</u> | <u>5</u> |
| <u>Increased depression, anxiety and distress</u> | <u>3</u> | <u>2</u> | <u>5</u> |
| <u>Loss of hope and motivation</u> | <u>2</u> | <u>1</u> | <u>3</u> |
| <u>Focus on primary needs</u> | <u>3</u> | <u>0</u> | <u>3</u> |
| <u>Idleness and antisocial behaviour</u> | <u>1</u> | <u>1</u> | <u>2</u> |
| <u>Stigmatisation</u> | <u>1</u> | <u>1</u> | <u>2</u> |

## Substance use issues

Substance abuse was a topic raised in all six focus groups, with participants concerned that young people were using alcohol, marijuana and other drugs as a coping mechanism during the pandemic, which led to addiction and poorer mental health (table 2). Participants noted that while some youth had experimented with substances before the pandemic, their use intensified during lockdowns due to stress, isolation, and lack of structured activities. As one participant from Malawi shared: *“People started using drugs just to forget everything—they think it helps, but it only makes things worse in the end” [M-FG-1].* A participant from Sudan noted: *“People are just smoking weed and taking something called ‘ice’ to numb themselves” [S-FG-2],* underscoring the extent to which substance use was seen as a form of emotional escape.

In both countries, marijuana and locally brewed alcohol were the most mentioned substances. Gender differences were observed, with male youth more often associated with public drug use and antisocial behaviours, while female participants were more likely to report alcohol use in private settings as a way to cope with emotional stress. Reasons for use included boredom, unemployment, the closure of schools, peer influence, and a desire to escape family pressures. Participants also linked substance use to broader social consequences, such as increased idleness, rising youth crime, and disengagement from school or community activities. In some cases, young people were described as becoming more withdrawn or aggressive, with substance use worsening their mental health and increasing the risk of long-term dependency.

## Economic instability

Economic downturns worsened by the COVID-19 pandemic led to widespread unemployment, sudden job losses, and overall financial instability, which exacerbated anxiety and depression among young people. In Sudan, participants described severe economic challenges, with even meeting basic needs becoming a daily struggle. As one participant explained: *“Even getting soap or bread became difficult— everything was expensive, and there was no work. We were just trying to survive each day*” *[S-FG-1].* Another participant highlighted the impact of unemployment, stating: *“Many people lost their jobs during COVID, especially young people. There was nothing to do, no income—it was very stressful” [M-FG-1].* These economic hardships left many participants feeling hopeless about their prospects. As one young person put it: *“There’s no future here anymore. If I had the chance, I would leave the country” [S-FG-2]. Another participant echoed this sense of prolonged despair, asking simply: “Till when will we struggle?” [S-FG-3]—*a reflection of growing fatigue and uncertainty about whether their situation would ever improve.

## Social isolation

Social isolation due to lockdowns and movement restrictions was a major challenge for young people across all focus groups. Participants shared how prolonged time at home, separation from friends and extended family, and the closure of schools and community spaces contributed to feelings of loneliness, boredom, and emotional disconnection. This lack of social interaction not only impacted their mental wellbeing but, for some, led to harmful coping mechanisms. As one participant shared: *“We were locked in for months. No school, no friends. You start to feel like no one cares, like you’re alone in everything” [S-FG-3].* In some cases, this isolation was linked to increased substance use, as young people sought ways to escape their thoughts or fill the emotional void. Others described feeling withdrawn and unmotivated, with the lack of routine and meaningful engagement deepening their sense of hopelessness

### Theme 2: Barriers to Accessing Mental Health Services

In this theme, participants spoke about different personal and societal issues that affected their ability to access mental health services or seek support from those around them (Table 3).

**Table 3:** Topics linked to ‘Barriers to Accessing Mental Health Services’.

|  | **<u>Sudan (3)</u>** | **<u>Malawi (3)</u>** | **<u>Total FGD (6)</u>** |
| --- | --- | --- | --- |
| <u>Stigma towards mental illness</u> | <u>3</u> | <u>2</u> | <u>5</u> |
| <u>Lack of awareness</u> | <u>3</u> | <u>2</u> | <u>5</u> |
| <u>Lack of knowledge about services</u> | <u>1</u> | <u>2</u> | <u>3</u> |
| <u>Unavailability of resources and affordability</u> | <u>2</u> | <u>1</u> | <u>3</u> |
| <u>Perceiving problem as not serious enough</u> | <u>1</u> | <u>1</u> | <u>2</u> |
| <u>Culture and toxic masculinity</u> | <u>1</u> | <u>1</u> | <u>2</u> |

## Stigma towards mental illness

Stigma surrounding mental illness emerged as a critical barrier to mental health care, perpetuating feelings of shame, fear, and social isolation. This issue was reported in five out of six focus groups, highlighting its pervasive impact across different contexts. Participants described how being associated with mental illness often led to social shame, discrimination, or being labelled as “mad”—labels that carried deep cultural weight and negative connotations. Such perceptions discouraged individuals from seeking support, even when they were in distress.

For example, one participant from Sudan explained, *“If you go to the clinic and say you have mental health issues, people think you’re crazy. They start treating you differently, even your own family” [S-FG-1].* This reflects how stigma can infiltrate not only the broader community but also intimate social circles, leading to a breakdown of trust and support.

Similarly, in Malawi, participants voiced concerns about judgment from both healthcare workers and the community. One participant shared, *“People will say you’re possessed or bewitched, not sick. So, you keep quiet because you don’t want everyone talking about you” [M-FG-2].* This fear of spiritual misinterpretation and gossip reinforces silence, making it harder for individuals to access appropriate care or even acknowledge their mental health needs.

## Lacking mental health awareness

A general lack of awareness about mental health and its symptoms was raised in five of the six focus groups. Participants described how mental health is not commonly discussed or understood, which often led individuals to overlook early signs of distress or misinterpret them. As a participant from Malawi shared: *“Most people don’t even know what depression is—they just think you’re being lazy or weak” [M-FG-1].* This limited understanding extended to knowledge about mental health services. In three focus groups, participants noted that even if someone recognised that they needed help, they often did not know where to go or how to access support. As one Sudanese participant put it: *“Even if you want to get help, you don’t know where to start. There’s no clear place for mental health” [S-FG-2].* A participant from Malawi echoed this sentiment: *“It’s hard to find help. There’s nowhere to go unless you have money or know someone who works in a hospital” [M-FG-2]*.

The lack of awareness and understanding also contributed to mental health not being taken seriously within their communities. Participants described a tendency for others to downplay emotional struggles, often attributing them to weakness or laziness. This dismissive attitude left some internalising the belief that their experiences were not valid or worthy of support. As one Sudanese participant explained: *“You tell someone you’re feeling stressed, and they say, ‘Just get over it.’ So, you start thinking maybe it’s nothing*” *[S-FG-3]*.

## Unavailability of resources and affordability

Three focus groups (two in Sudan, one in Malawi) highlighted practical barriers such as the high cost of services, lack of actual mental health facilities, and a shortage of trained mental health professionals. These limitations made it nearly impossible for some young people to access formal mental health care.

As a young person from Malawi shared: *“Mental health care is expensive, and even if you have money, there are no proper clinics near us” [M-FG-2].* A participant from Sudan similarly explained: *“There are no psychologists or counsellors here. And if there are, it’s only in big hospitals and it costs too much” [S-FG-1]*.

In the absence of accessible professional support, many participants said they turned to informal networks—speaking to friends, family, or religious leaders—for emotional support. However, some noted that this was not always helpful or appropriate. As one Sudanese participant said: *“Sometimes you talk to your friends, but they don’t know what to say. They just tell you to pray or stop thinking too much” [S-FG-2].* Others reported doing nothing at all, choosing instead to suppress their feelings due to stigma or lack of options. A few participants also connected the unaffordability of care to the rise in unhealthy coping strategies, particularly substance use. With nowhere else to turn, some young people resorted to drugs or alcohol to manage their distress. As one participant from Malawi said: *“People know they need help, but they can’t afford it, so they smoke or drink to forget” [M-FG-3].* These responses suggest that systemic barriers to accessing mental health care not only leave emotional needs unmet but may also contribute to worsening outcomes through maladaptive coping.

## Culture and Toxic Masculinity

Cultural expectations — particularly around masculinity — were cited in two focus groups as a barrier to seeking help for mental health issues. Participants described how societal norms discourage men from expressing vulnerability or admitting emotional struggles. A male participant from Malawi reflected: *“As a man, you’re supposed to be strong all the time. If you say you’re struggling, people will laugh or call you weak” [M-FG-2].* Similar sentiments were echoed in Sudan, where a female participant observed: “*Men here feel like they always have to hide their emotions. If they cry or say they’re stressed, people say they’re not real men*” *[S-FG-1].* These perceptions of masculinity created pressure to suppress emotions and avoid seeking support, even when it was clearly needed.

While much of the discussion focused on expectations placed on men, some participants also reflected on gender differences in emotional expression. A few female participants noted that women were somewhat more “allowed” to express feelings or seek help without being judged as weak. As one Sudanese participant shared: *“At least as a girl, if you cry or say you’re feeling low, people understand. But if a guy does it, it’s shameful” [S-FG-2].* These gendered norms contributed to unequal barriers, with young men in particular facing stigma when confronting or disclosing mental health struggles.

### Theme 3: Ways to Access Mental Health Support

Despite the challenges impacting mental health and the barriers to accessing support raised by this cohort of young people during their FGDs, there were also examples shared of their experiences when they accessed existing mental health support along with ideas to improve service provision for young people (Table 4).

**Table 4-:** Topics linked to ‘Ways to Access Mental Health Support’.

|  | **<u>Sudan (3)</u>** | **<u>Malawi (3)</u>** | **<u>Total FGD (6)</u>** |
| --- | --- | --- | --- |
| <u>Support from educational system</u> | <u>1</u> | <u>3</u> | <u>4</u> |
| <u>Professional counselling</u> | <u>2</u> | <u>2</u> | <u>4</u> |
| <u>Group and club discussions</u> | <u>2</u> | <u>1</u> | <u>3</u> |
| <u>Support from family and friends</u> | <u>3</u> | <u>0</u> | <u>3</u> |
| <u>Raising awareness on mental health</u> | <u>3</u> | <u>0</u> | <u>3</u> |
| <u>Hospital and rehab centres</u> | <u>1</u> | <u>1</u> | <u>2</u> |
| <u>Hotline for mental health services</u> | <u>2</u> | <u>0</u> | <u>2</u> |

## Support from community-based structures

Participants from four FGDs (three in Malawi, one in Sudan) discussed how schools and universities could serve as important settings for promoting mental health awareness and support. Participants emphasised the need for educational institutions to offer structured counselling, regular emotional check-ins, or safe, stigma-free spaces for students to talk about their mental health needs. A participant from Malawi shared their experience of receiving support at school: *“At school, at least we had someone to talk to — a teacher or a counsellor who would check in if you looked stressed” [M-FG-2].* This example highlighted the role that educational settings can play as a first point of contact for mental health support.

In Sudan, while few participants had direct experience with school-based services, several expressed a strong desire for such support to be available. One participant remarked: *“We don’t have anything like counsellors in schools here, but it would help if there was someone to talk to, especially for students who are struggling” [S-FG-1].* The absence of structured support in schools was seen as a missed opportunity, particularly as academic stress and emotional pressure were commonly reported.

Beyond schools, discussions during community youth groups and clubs were described as helpful ways to share experiences and reduce feelings of isolation. This was highlighted in three focus groups (two in Sudan, one in Malawi). Peer support in these more informal but structured environments was viewed as empowering and effective in helping to normalise conversations around mental health. As a Sudanese participant explained: *“In our youth group, we talk about everything—even depression. Just knowing others feel the same helps a lot” [S-FG-2].* These groups provided safe, collective spaces where young people could open up without fear of judgment, particularly in contexts where formal services were lacking or inaccessible.

## Experiences of professional support

Professional mental health services were recognised as a key form of support in four focus groups (two in each country). Although access was often limited, the few participants who had accessed professional support valued the role of trained counsellors in helping them understand and manage their emotional struggles. One Sudanese participant reflected, *“I had one session with a counsellor, and even that helped me feel like I wasn’t alone or crazy” [S-FG2]* This quote illustrates the profound impact that even a single counselling session can have on young people feeling isolated in their struggles.

In two FGDs (one from each country), hospitals and rehabilitation centres were acknowledged as essential—but often under-resourced—facilities. Participants shared both gratitude for and frustration with the limitations of these services. A participant from Malawi noted, *“There’s a rehab center in the city, but it’s far and expensive. Still, it’s the only place that understands mental health properly” [M-FG-1]* This quote underscores the challenges faced by individuals seeking more formal, structured mental health care.

## Support from Family and Friends

Support from close social networks was seen as crucial across all three focus groups in Sudan. Young people described relying on trusted family members or friends for emotional release, comfort, and guidance during times of distress. One Sudanese participant shared: *“Sometimes just talking to my sister or a close friend made things easier. They didn’t judge me, they just listened” [S-FG-1].* This illustrated the vital role that informal support systems played in providing immediate, non-judgmental care, particularly in the absence of professional services.

Another participant described turning to their mother when overwhelmed by anxiety during the pandemic: *“There were days I couldn’t sleep because I was so stressed. My mother would sit with me, talk to me, and pray with me. It made me feel less alone” [S-FG-2].* In some cases, the support wasn’t just emotional but also practical. One young person recalled: *“When I was feeling down, my cousin made sure I came out of the house, ate something, or joined them for a walk. They didn’t let me isolate” [S-FG-3]*.

These examples highlight how emotional closeness, small acts of care, and being listened to without judgement contributed to a sense of relief and connection. Interestingly, no participants from Malawi shared experiences of seeking or receiving similar support from close family or friends, which may reflect differences in family dynamics, stigma, or communication norms around mental health across the two countries.

## Ideas for improving youth focused service provision

Participants across multiple focus groups suggested two key ways to improve mental health support for young people. The first was improving mental health literacy at all levels of the community. This was discussed in at least five focus groups — three in Sudan and two in Malawi. Participants across both contexts highlighted the need to raise awareness, reduce stigma, and normalise mental health conversations. Suggestions included community-based education campaigns, integrating mental health into school curricula, and using mass media (such as radio and television) to spread positive messages. As one participant from Malawi put it*: “We need more people talking about this—on the radio, in schools, in churches—so it doesn’t feel like a secret” [M-FG-2].* Another Malawian participant similarly noted: “*If we were taught about this in school, it would be easier to understand when you’re not okay. Right now, most people don’t even know what mental health is” [M-FG-3]*.

The second suggested improvement was the introduction or expansion of mental health hotlines, which came up in two Sudanese focus groups and was also mentioned in Malawi. While such services were often unavailable or unreliable, participants saw them as a potentially valuable, low-barrier tool for immediate support. A Sudanese participant shared: “*A hotline would be good—sometimes you just need someone to talk to at night when things feel too heavy” [S-FG-1].* Participants pointed to several reasons why hotlines would be helpful: they offer anonymity, which reduces fear of being judged; they are convenient, especially for young people without access to transport; and they could be free or low-cost, removing financial barriers that often prevent access to professional care. One participant explained: *“It’s easier to call a number than to go somewhere and be seen. If it’s free, more people would use it” [S-FG-3]*.

## Discussion

This study aimed to explore the impact of the COVID-19 pandemic on the mental health of young people in Malawi and Sudan through qualitative focus group discussions. The findings underscore the profound and multifaceted effects the pandemic had on this demographic, highlighting a complex interplay of psychological, social, and economic stressors.

Themes identified from the focus groups reveal meaningful increases in anxiety, depression, and stress experienced and perceived participants. These effects were compounded by economic instability and prolonged social isolation. Of particular concern is the reported rise in substance abuse among young people, used as a coping mechanism to manage pandemic-induced stress and uncertainty. This aligns with existing literature showing that during public health crises, substance use often increases as a maladaptive strategy to handle psychological distress [1].

Economic instability was a prominent source of stress, especially for young people in transitional life stages—either entering the workforce or still engaged in education. The loss of employment opportunities and disruptions to academic routines, including the shift to online learning, created additional psychological burdens. This is consistent with prior research indicating that economic downturns can severely affect young people’s mental health, especially when paired with limited social and financial support systems [2].

Social isolation further exacerbated mental health challenges. Lockdown measures disrupted daily routines and removed key sources of emotional support, such as peer interactions and community activities. The crucial role of social connectedness in promoting mental well-being is well-established, and its absence during the pandemic led many participants to report feelings of loneliness, detachment, and hopelessness [3].

A significant barrier to mental health recovery is the ongoing stigma surrounding mental health services. This stigma—prevalent across many African populations and low– and middle-income countries (LMICs) more broadly—continues to deter individuals from seeking help [18]. Cultural norms, low mental health literacy, and limited access to affordable and culturally appropriate services contribute to this reluctance. Participants in both countries described mental health challenges as being dismissed or misunderstood in their communities, which worsens the isolation and distress they experience. Similar trends have been documented in studies from Asia and South America [19,20], emphasizing the global nature of this challenge in LMIC contexts.

### Study Strengths

One key strength of this study is its use of qualitative focus groups to gather rich, in-depth insights directly from young people from two under-researched LMIC contexts—Malawi and Sudan. This approach allowed for a nuanced understanding of how the pandemic has impacted mental health, going beyond statistics to explore lived experiences, emotions, and coping strategies. The inclusion of participants from both urban and rural settings further enhances the diversity views within findings. Additionally, the study contributes to the growing body of literature on youth mental health in LMICs, offering valuable perspectives that are often overlooked in global discussions.

### Study limitations

Despite its contributions, the study has several limitations. Firstly, the use of focus groups may have introduced social desirability bias, as participants might have been hesitant to openly discuss sensitive issues such as substance abuse or mental illness in a group setting. Secondly, the online or socially distanced format of the discussions—necessitated by pandemic restrictions—may have excluded young people affected by digital poverty i.e. with no access to devices or internet data. This is a common challenge in the geographical setting of this study, evidenced by the five individuals who were recruited to the study but were subsequently unable to join the Zoom platform. Furthermore, connectivity issues may delay or halt speech during online discussions which can affect the depth and spontaneity of participant responses.

## Conclusion

This study provides valuable insights into the mental health challenges faced by young people in Malawi and Sudan during the COVID-19 pandemic. Findings highlight the urgent need for targeted interventions to address the unique challenges identified, including economic instability, social isolation, substance abuse, and cultural stigma that intersect to negatively impact individuals’ mental health.

To mitigate the negative impact of pandemics on youth mental health, several strategies are recommended. First, promoting positive attitudes towards mental health and reducing stigma through public awareness campaigns can improve understanding of mental health at grassroots levels and encourage more young people to seek help. These campaigns need to include tackling the issue of substance use, so that people are aware that taking alcohol or drugs is a maladaptive coping mechanism for mental distress, particularly in the context of the added pandemic stressors. Second, improving access to mental healthcare services by investing in infrastructure and training more mental health professionals is essential. Thirdly, decision makers need to acknowledge the important role social connections and economic stability play in the mental wellbeing of young people. Therefore, during future pandemics, efforts to maintain these connections through virtual platforms and community support initiatives can help provide much-needed social interaction and support to mitigate mental illness among this population. Additionally, providing economic support and opportunities for young people can alleviate some of the financial stress and uncertainty they face.

## Data Availability

All relevant data are within the manuscript and its Supporting Information files.

## Acknowledgements

The authors would like to thank all their participants involved in this study, especially during a very challenging time.

